# Genetic, Cellular, and Connectomic Characterization of the Adult Human Brain Regions Commonly Plagued by Glioma

**DOI:** 10.1101/2020.02.18.20018531

**Authors:** Ayan S. Mandal, Rafael Romero-Garcia, Michael G. Hart, John Suckling

## Abstract

A better understanding of the nonrandom localization patterns of gliomas across the brain could lend clues to the origins of these types of tumors. Following hypotheses derived from prior research into neuropsychiatric disease and cancer, gliomas may be expected to localize to brain regions characterized by hubness, stem-like cells, and transcription of genetic drivers of gliomagenesis. We combined neuroimaging data from 335 adult patients with high- and low-grade glioma to form a replicable tumor frequency map. Using this map, we demonstrated that glioma frequency is elevated in association cortex and correlated with multiple graph-theoretical metrics of high functional connectedness. Brain regions populated with stem-like cells also exhibited a high glioma frequency. Furthermore, gliomas were localized to brain regions enriched with the expression of genes associated with chromatin organization and synaptic signaling. Finally, a regression model incorporating connectomic, cellular, and genetic factors explained 58% of the variance in glioma frequency. Our findings illustrate how factors of diverse scale, from genetic to connectomic, can independently influence the anatomic localization of oncogenesis.

## Introduction

Tumor location represents one of the most important prognostic factors for patients suffering from primary brain cancers^1,2^, yet little is known about the mechanisms that determine the spatial distribution of gliomas across the brain.

The importance of glioma location for diagnosis and treatment has been recognized since Percival Bailey and Harvey Cushing’s seminal classification of brain tumors in the early 20^th^ century^3^. Whilst brain imaging, primarily MRI, plays an important and routine role in diagnosis and treatment of brain tumors, there has been little quantitative mapping of their distribution at a population level. Comprehensive modeling of the key factors involved in determining why gliomas might be heterogeneously distributed across the brain could shed light on the origins of these tumors, and consequently inform treatment targets. Three general hypotheses for the spatial distribution of gliomas include a “connectomic hypothesis”, a “cellular hypothesis”, and a “genetic hypothesis”, and each is now considered in turn.

The connectomic hypothesis posits that highly connected brain regions, known as hubs, are especially vulnerable to disorders, such as oncogenesis, due to the metabolic costliness of maintaining many connections^4,5^. The term “connectome” was originally conceived by analogy to the term “genome”, and refers to the collection of all connections, anatomical or functional, in the brain^6^. The connectome can be defined at the microscale, in which case the connections represent synaptic links between neurons, or at the macroscale, where the connections can represent anatomical white matter pathways (structural connectome), or correlations in neuronal activity (functional connectome) between brain regions^7^. A foundational finding of network neuroscience is that the connectomes of many different species, across multiple scales, possess a small-world architecture^8^. In other words, they are composed mostly of short distance connections between neighboring nodes (brain regions or neurons), but with a few long-distance connections between distant nodes. The nodes from which originate many of the short and long distance connections are crucial for efficient communication across the network, and these are the hubs^9^. Brain hubs are believed to be “costly” due to the metabolic demand of maintaining many connections^10^, a factor that makes these regions vulnerable to disease^11–14^. Long distance axonal connections for instance, are physiologically expensive to maintain since proteins in the neuron’s presynaptic terminal must be produced in the nucleus, and thus travel the full distance of the axon to reach their target. This factor contributes to the vulnerability of upper motor neurons to degeneration^15^. In a similar way, long distance connections important for the construction of large-scale cortical networks also pose a challenge for glial cells (in particular, oligodendrocytes) to support the requisite axonal tracts^16^. Furthermore, brain hubs also receive many connections, and therefore are populated with many synapses which impose metabolic demand upon supporting astrocytes^17^. Metabolic demands on the glial cells of hub regions could contribute to elevated cell turnover, enhancing the likelihood of a cell acquiring an oncogenic mutation during mitosis^18^. Metabolic stress could also contribute to oncogenesis via enhanced production of mutagenic reactive oxygen species^19^. For these reasons, one may expect gliomas to localize to hubs of the brain connectome.

With their shared dedifferentiated and proliferative nature, the commonalities between stem cells and cancer cells have not gone unnoticed among cancer biologists. These commonalities form the basis of the stem cell hypothesis of cancer, which maintains that cancers tend to originate from normal stem and stem-like cells in the body^20–22^. When applied to adult glioma, this hypothesis points to two clear suspects as possible cells-of-origin: neural stem cells (NSCs) and oligodendrocyte precursor cells (OPCs)^21,22^. Neither are randomly distributed throughout the brain, and therefore their specific localization patterns have been hypothesized to play a role in determining the nonrandom distribution of gliomas^23,24^. The notion that neural stem cells exist and continue to proliferate in the adult human brain is relatively new and historically controversial, but a consensus has arisen that they can be found in at least two locations: the subgranular zone of the dentate gyrus of the hippocampus, and the subventricular zone ^22,25^. Rodent work has demonstrated that OPCs are widely distributed throughout the mammalian brain^26^. The patterning of OPCs in the adult human brain is unclear, but could be estimated by utilizing brain-wide maps of gene expression patterns^27,28^.

Adult gliomagenesis is the result of glial cells acquiring a series of somatic mutations which trigger uncontrollable cell proliferation^30,31^. Recent research has demonstrated that tumor location is influenced by the genetic aberrations that guide the development of tumors^23,29^.

Gliomas may be expected to localize to brain regions where the genetic risk factors for the disease are normatively expressed. Furthermore, consequential to the connectomic and cellular hypotheses, it may be expected that brain regions frequented by glioma are enriched with the expression of genes associated with cell proliferation or metabolically intensive processes required for long distance neuronal signaling.

In this study, we tested these three hypotheses by examining the connectomic, cellular, and genetic correlates of brain regions commonly plagued by glioma. We began by deriving a replicable tumor frequency map from neuroimaging data of 335 adult patients with high- and low-grade glioma. Using this map, we compared glioma distributions across canonical subnetworks and correlated them with hub measures calculated from averaged functional connectivity data from a large number of healthy individuals. Then, we determined if glioma frequency was elevated among brain regions expected to be enriched with NSCs and OPCs.

Next, we conducted a transcriptomic analysis to find genes with spatial expression patterns that followed the observed glioma distribution. Finally, we combined all these factors of glioma distribution into a single regression model to explore the putative inter-relationships of predictors of glioma frequency.

## Results

### Anatomical mapping of glioma distribution

We initially constructed a map of glioma distribution from aligned masks of tumor volume across 335 high- and low-grade glioma patients. This tumor frequency map displayed a hemispherically symmetric, but heterogeneous spatial distribution (Figure 1A). Consistent with prior reports, gliomas were rare in the occipital lobe, but relatively common in insular cortex (Figure 1B; Supplementary Table 1). Tumor frequency distributions were replicable across independent, randomly assigned subsets of half of the images (Groups 1 and 2) with an inter-regional correlation of r=0.83 (95% CI: r=0.70-0.93). Replicability of subsequent analyses was tested with Group 1 and Group 2 tumor frequency maps (see Supplementary Information).

**Figure 1.**
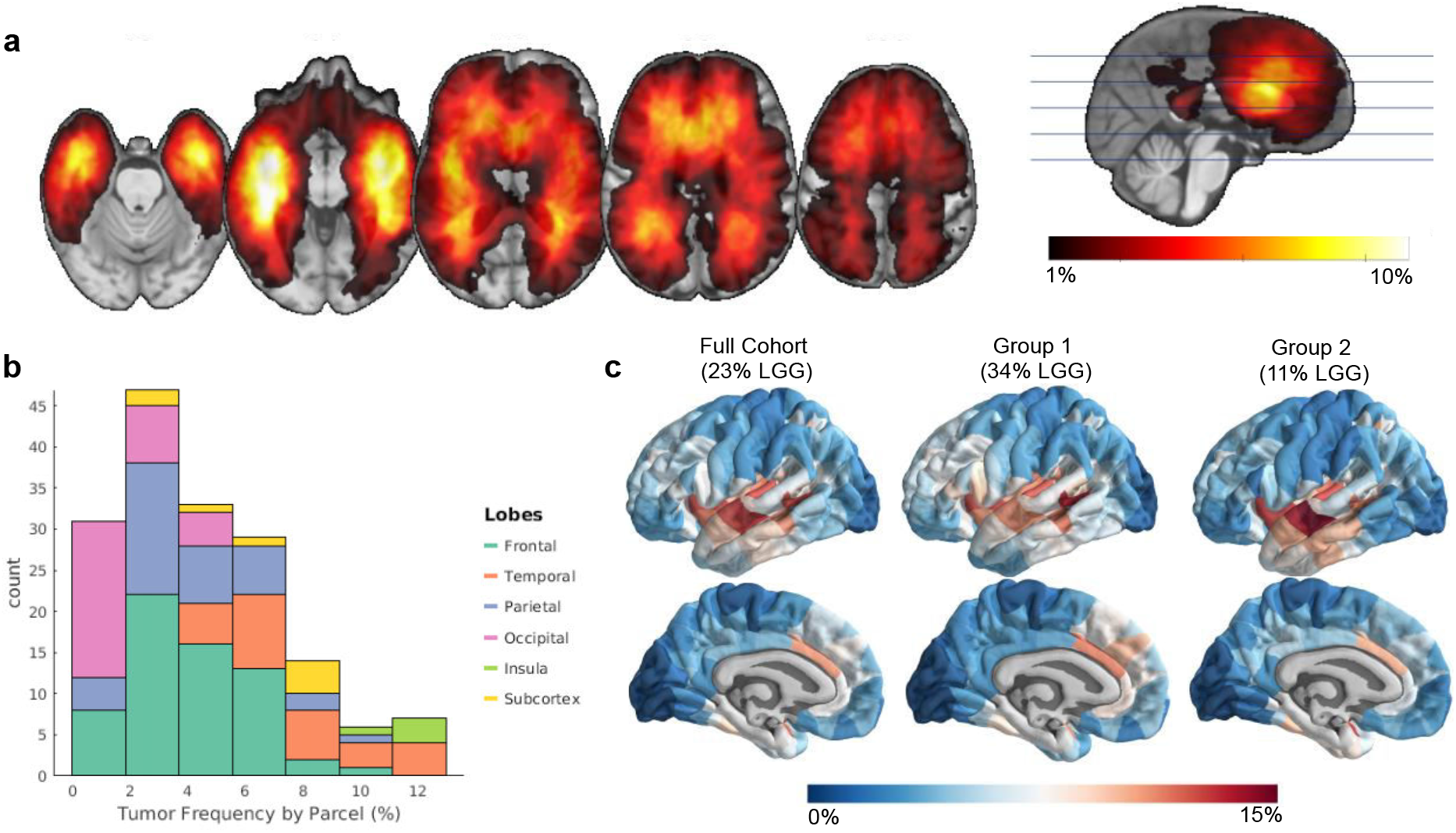
Non-random spatial distribution of gliomas. A. Tumor frequency map derived from lesion masks from 335 patients with high- and low-grade glioma. B. Glioma frequency by common anatomic subdivisions. C. Glioma frequency represented at a parcel-level. Internal replicability of glioma frequency tested by constructing two independent maps from even splits of the cohort, where the first comprised of ∼34% low-grade gliomas and the other of ∼11% low-grade gliomas.

Tumor frequency was compared across canonical, large-scale functional networks and primary versus association cortex. Association regions responsible for consolidating information across multiple sensory modalities showed higher tumor prevalence (average voxel: 4.57%) than visual and somatosensory primary cortices which had the lowest tumor frequency (2.45%), particularly in the visual cortex (1.56%; Figure 2A and 2B).

**Figure 2.**
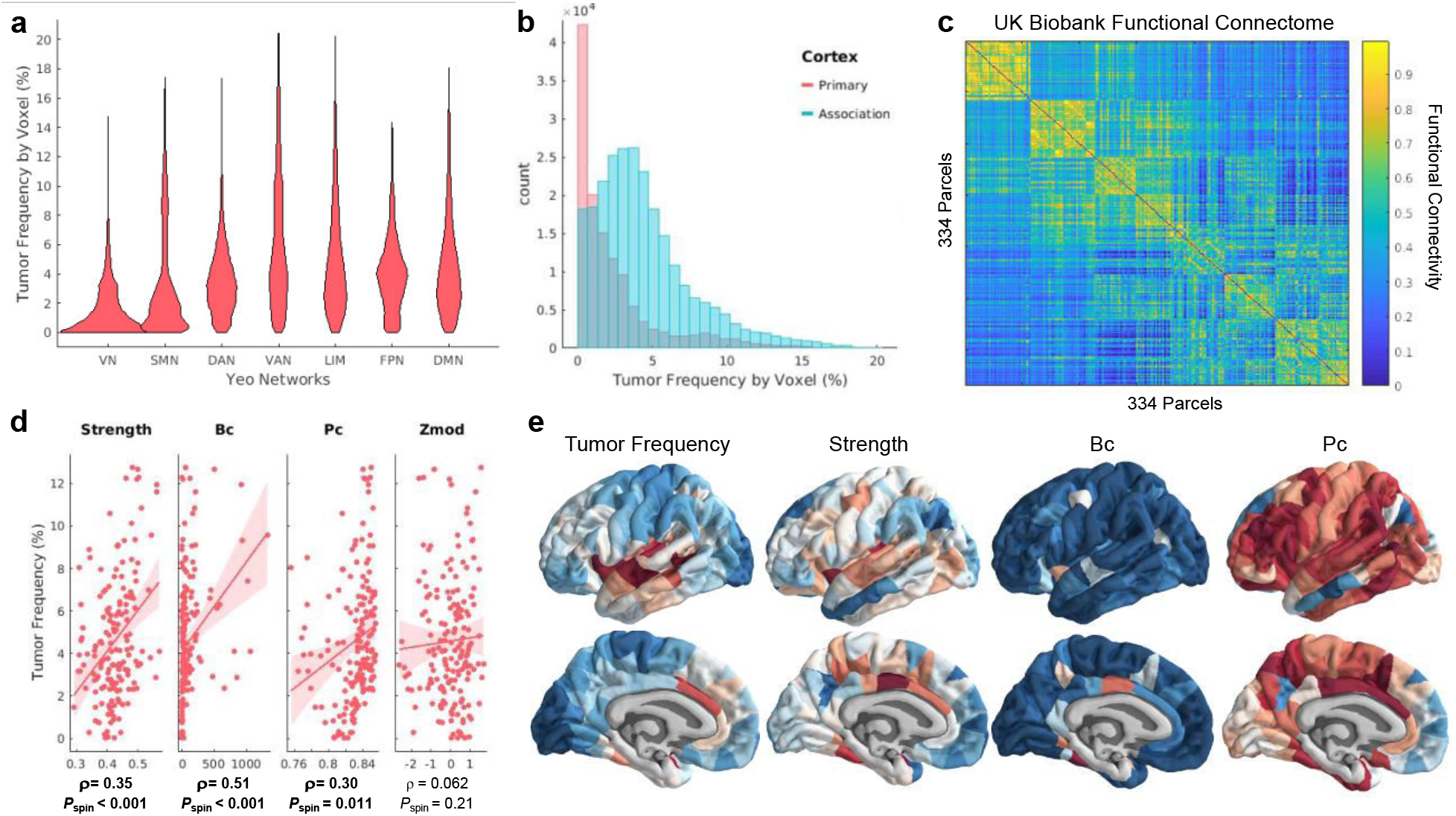
Gliomas localize to connector hubs of the brain. A. Violin plot comparing glioma frequency distributions across canonical subnetworks. B. Histogram comparing glioma frequency distribution across primary versus association cortex. C. Functional connectome calculated from resting state functional scans of over 4000 UK BioBank participants. Nodes in the network are organized according to their affiliation with different canonical subnetworks. D. Correlations between glioma frequency and hub measures calculated from the functional connectome. E. Visualization of glioma frequency and functional hub measures on the cortical surface.

### Gliomas localize to hubs of high connectivity and centrality

Graph theory measures were calculated from the mean functional connectome derived from resting state fMRI scans of over 4000 UK BioBank participants, and then compared with glioma frequency. The connectome was first organized into seven communities of interconnected nodes based on their overlap with previously defined large-scale functional networks^32^ resulting in a neat modular organization, with strong connections within modules and sparse connections between modules (Figure 2C). Graph theory measures of hubness were then calculated, measuring properties such as connectivity with neighboring nodes, involvement in shortest paths across the network, connectivity to nodes in different modules, and within-module connectivity (see *Methods* for how these measures were defined and selected). The inter-regional correlation between measures of hubness and glioma frequency was tested for significance by comparison to spatially contiguous null models (Figure 2D).

Glioma frequency strongly correlated with the simplest measure of hubness, nodal strength (ρ = 0.34; *P*_spin_ = 0.00055), which aggregates the weights of a node’s immediate connections. Glioma frequency was also significantly correlated with betweenness centrality (ρ = 0.51; *P*_spin_ = 0.0002) and with a measure of connectivity to diverse communities, the participation coefficient (ρ = 0.30; *P*_spin_ = 0.011). Connectivity within a community, measured by Z-score modularity, did not relate with glioma frequency (ρ = 0.062; *P*_spin_ = 0.21; Figure 2D). This profile of connectivity measures was most consistent with that of connector hubs that link together multiple sub-networks.

### Glioma frequency is elevated in areas with populations of stem-like brain cells

We tested the hypothesis that brain regions enriched with NSCs were more likely to coincide with loci of high frequency of gliomas. Mean tumor frequency was calculated from the hippocampus and the caudate (Figure 3A), regions which best approximate the locations of the only known sources of NSCs in the adult human brain. Tumor frequency across bilateral hippocampus and caudate were averaged, and compared against a null distribution of 10000 different pairs of randomly selected parcels within our 334-region parcellation scheme. Glioma frequency was observed to be significantly higher in these two regions compared to the null distribution (*p* = 0.0315; Figure 3B).

**Figure 3.**
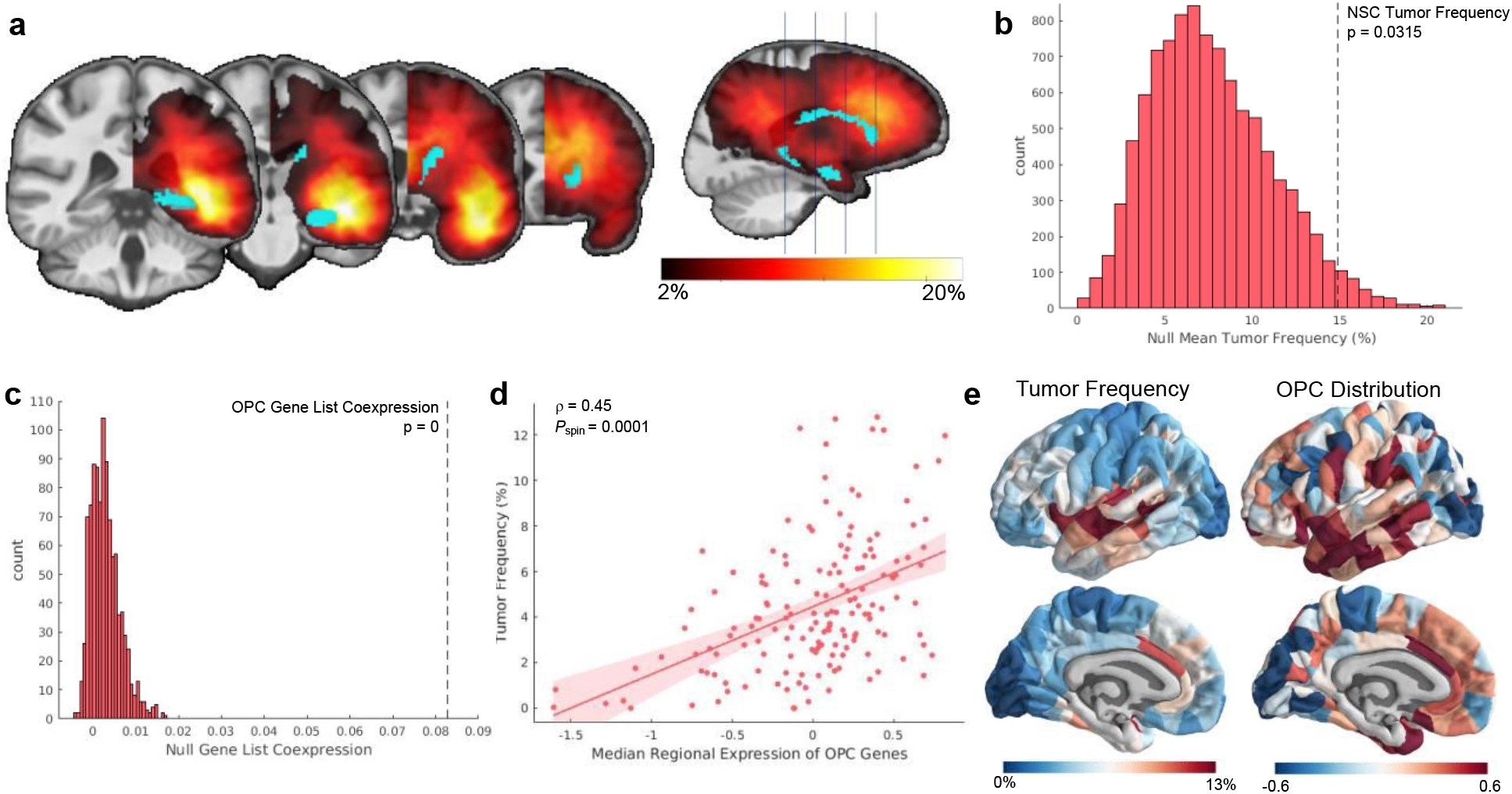
Gliomas localize to brain regions enriched with stem-like cells. A. Visualization of the parcel masks representing the hippocampus and caudate superimposed on the mirrored tumor frequency map. B. Average tumor frequency across the hippocampus and caudate (represented as the dotted black line) compared to a distribution of average tumor frequency across 10000 sets of two randomly chosen parcels. C. Co-expression among genes within the OPC gene list compared to co-expression among 10000 identically-sized sets of genes. D. Correlation between OPC distribution across cortex and glioma frequency (ρ = 0.45; *P*_spin_ = 0.0001). E. Visualization of glioma frequency and OPC distribution on the cortical surface.

Next, we tested the spatial correspondence of glioma distribution with the patterning of OPCs, which are also hypothesized to be cells-of-origin for glioma. OPC distribution was estimated from the expression of genetic markers of OPC identity using post-mortem the microarray data of the Allen Human Brain Atlas (AHBA; www.brain-map.org). The list of genetic markers for OPC’s co-expressed significantly (Figure 3C) confirmed that median expression across this gene list represents a spatially specific phenotype. This estimate of OPC patterning correlated significantly with glioma frequency (Figure 3D; ρ = 0.45; *P*_spin_ = 0.0001).

### Transcriptomic correlates of glioma frequency

We used partial least squares (PLS) regression to relate the spatial transcription patterns of 20647 genes with tumor frequency at 2748 cortical and subcortical locations where gene expression was assessed in postmortem adult human brain tissue (Figure 4A,B). The first two components of the PLS (PLS1 and PLS2) explained 19% and 18% of the tumor frequency variance, respectively (Supplementary Figure 3A), more than expected by chance (Permutation test; *p* < 0.001; Supplementary Figure 3B).

**Figure 4.**
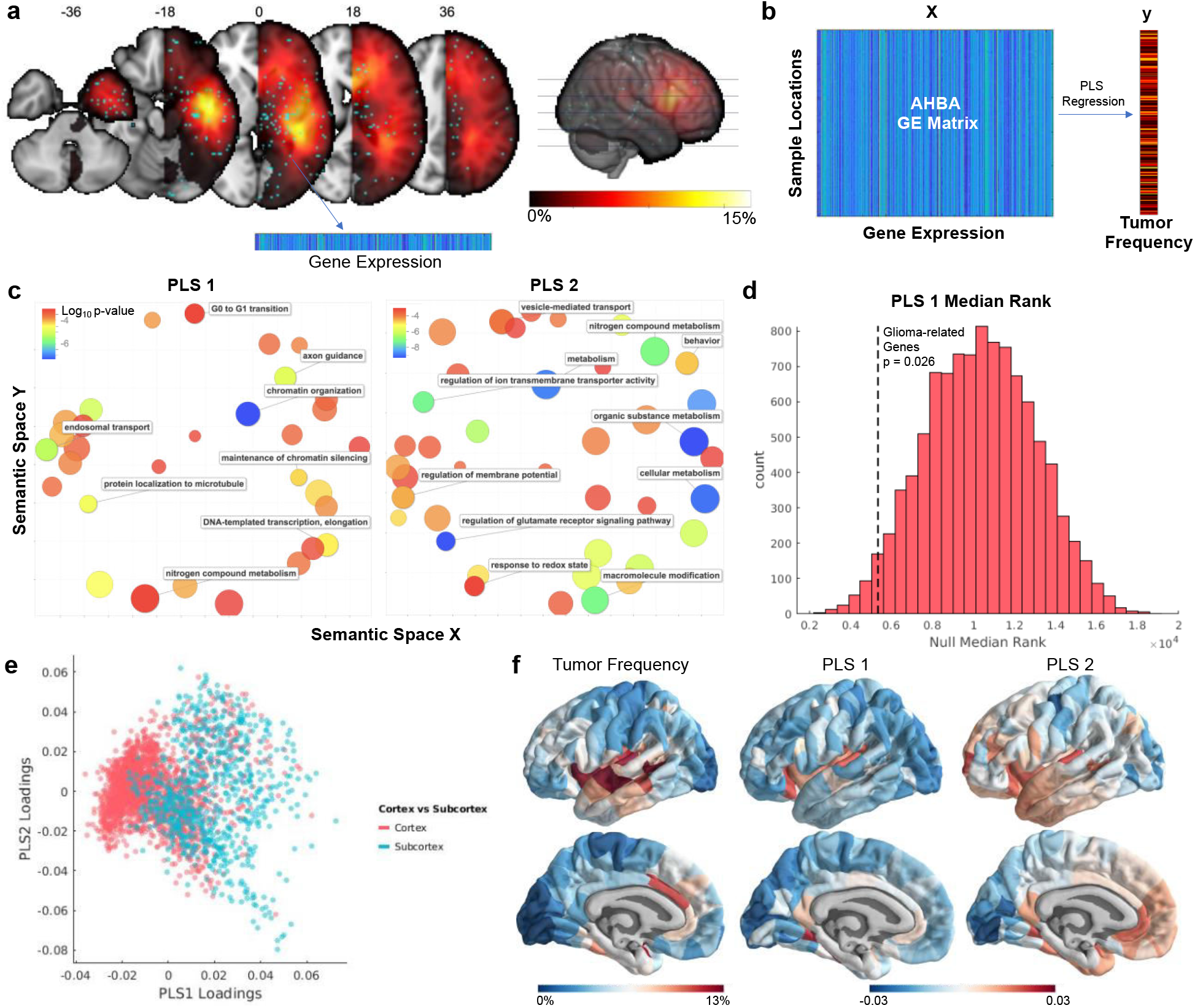
Transcriptomic correlates of glioma frequency. A. Alignment of AHBA sample locations to the tumor frequency map. B. Illustrative flowchart of the statistical analysis relating normative spatial gene expression patterns to glioma frequency. C. Gene ontology terms associated with two partial least squares components (PLS1 and PLS2) that related gene expression with glioma frequency. D. Median rank of 13 genes commonly altered in glioma compared to null distribution of median ranks. E. AHBA samples plotted by PLS1 loadings, PLS2 loadings, and cortex versus subcortex. F. Visualization of glioma frequency, PLS1 loadings, and PLS2 loadings on the cortical surface. PLS loadings from samples were assigned to parcels via a nearest neighbor mapping.

Bootstrapping was performed on PLS weights resulting in Z statistics for each gene corresponding to the PLS1 and PLS2 ranking (Supplementary Figure 3C). The ranked gene lists were entered into a gene ontology (GOrilla; http://cbl-gorilla.cs.technion.ac.il/). Genes corresponding to PLS1 were related to biological processes such as chromatin organization, endosomal transport, and G0 to G1 transition. Genes corresponding to PLS2 were related to a broad set of metabolic processes along with many components of synaptic transmission (Figure 4C). PLS1 was also found to be significantly enriched for genetic drivers of gliomagenesis (*p* = 0.026; Figure 4D). PLS2 was not significantly enriched for this set of genes (*p* = 0.75).

PLS1 was more highly loaded onto the subcortex relative to the cortex (Figure 4E). PLS loadings for each AHBA sample were mapped to their nearest brain region for visualization on the cortical surface (Figure 4F).

### Connectomic, cellular, and genetic contributions to glioma frequency are independent

Finally, we sought to reveal the interrelations between the connectomic, cellular, and genetic contributions to glioma distribution uncovered in the study. A multiple linear regression model was constructed, with factors of nodal strength, OPC distribution, PLS1 loadings, and PLS2 loadings (Figure 5A). NSC distribution was not included in the model because this measure could not be quantified at each brain parcel. First, we tested a model to determine if there were interaction effects between connectomic (nodal strength), cellular (OPC distribution), and genetic (PLS1 and PLS2 loadings) factors. None of the interaction effects were significant. The model without interaction effects explained approximately 58% of the variance in glioma frequency (F(4,162) = 59.3; *p* = 9.37 × 10^−31^; Adjusted R^2^ = 0.584; Figure 5B,C). All individual factors significantly predicted tumor frequency variance (Table 1; Figure 5D). Because of the unequal mapping of AHBA samples to cortical versus subcortical regions, the PLS2 component, which is more highly represented across cortex, explained more of the variance in tumor frequency than PLS1 once projected onto the anatomy. It is also worth noting that the amount of variance explained by the PLS factors was inflated by construction, due to the large number of input variables and the design of the technique which results in maximizing covariance^33^.

**Table 1.** Results of multiple linear regression model predicting tumor frequency.

| Predictor | Beta Value | Standard Error | T Statistic | Percent Explained | Spin Test Corrected P |
| --- | --- | --- | --- | --- | --- |

|  |  |  |  | Variance | Value |
| --- | --- | --- | --- | --- | --- |
| **Intercept** | 3.2e-16 | 0.0499 | 6.37e-15 | NA | NA |
| **Nodal strength** | 0.202 | 0.0543 | 3.72 | 7.86% | 0.0010 |
| **OPC distribution** | 0.206 | 0.0566 | 3.64 | 7.58% | 0.0040 |
| **PLS1 Loadings** | 0.210 | 0.0594 | 3.54 | 7.17% | 0.0052 |
| **PLS2 Loadings** | 0.480 | 0.0577 | 8.32 | 29.9% | 0 |

**Table 2.** List of glioma-related genes tested for enrichment among transcriptomic correlates of glioma distribution. These genes were selected from a recent review of molecular genetic markers of adult glioma subtypes^8^.

| Glioma-related genes |  |
| --- | --- |
| *IDH1* | *TP53* |
| *IDH2* | *NF1* |
| *TERT* | *MDM2* |
| *ATRX* | *PIK3CA* |
| *EGFR* | *FUBP1* |
| *CDKN2A* | *NOTCH1* |
| *CDKN2B* | *PDGFRA1* |
| *PTEN* | *RIS* |
| *RIK* | *PI3K* |

**Figure 5.**
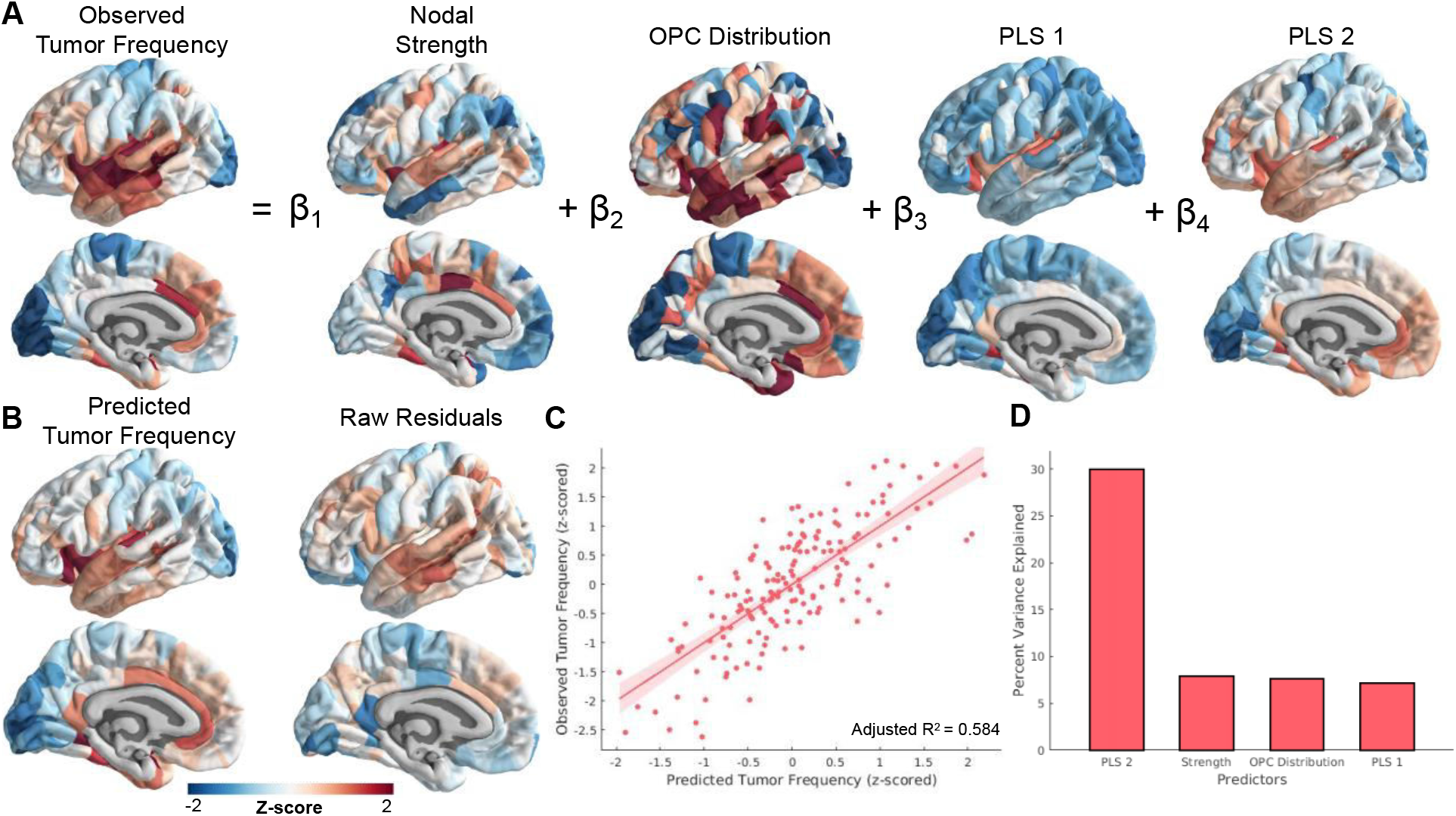
Multiple linear regression model relating connectomic, cellular, and transcriptomic factors with glioma distribution. A. Schematic of the multiple linear regression model. Intercept and error terms are not displayed. B. Fitted values and residuals of glioma distribution model. C. Scatter plot of predicted versus observed tumor frequency values. D. Percent of variance explained by each individual predictor of tumor frequency. These values were calculated using the partial correlation coefficient between each measure and tumor frequency.

## Discussion

In this study, we examined the network, cellular, and transcriptomic correlates of brain regions commonly frequented by glioma to test specific hypotheses regarding gliomagenesis. We found that gliomas were most common in association cortex and connector hub regions. Elevated glioma frequency was observed in brain regions expected to be populated by NSCs and OPCs. Finally, we determined that glioma distribution correlated with the spatial transcription patterns of genes related to metabolic activity, synaptic signaling, and gliomagenesis. These findings support the predictions of network neuroscience and cancer theory, and establish links between concepts from these two frameworks to characterize the spatial distribution of adult gliomas.

An extensive body of work has demonstrated the utility of network models in predicting the spread of disease^34–36^ as well as the vulnerability of particular brain regions to disease^11,12,14^. In this work, we used network models to demonstrate for the first time that functional hub regions of the brain are vulnerable to the concentration of gliomas. In particular, gliomas appear to localize to brain regions expected to play the role of connector hubs, nodes that link diverse cognitive subsystems with one another, as opposed to provincial hubs, which integrate communication within their own subsystems^9^. This suggests that the brain regions which facilitate long distance connections across the cortex are especially vulnerable to oncogenesis, consistent with our hypothesis that the high metabolic cost of such connections influences glioma risk^10^. Alternatively, the results can be interpreted as reflecting a higher likelihood for tumor infiltration of hub regions. Gliomas are known to migrate throughout the brain via blood vessels and white matter tracts, which contributes to the poor prognosis of glioblastoma multiforme. Here we consider only the location of the tumor during the pre-operative scan, which could represent either the tumor origin, or to where it spread during the progression of the disease. Although networks were constructed from functional MRI and not white matter tracts along which tumors are known to infiltrate^37^, recent work on activity-dependent glioma migration suggests that tumors could preferentially invade functional hubs^38^. Due to their high centrality, hubs are, by definition, likely to be encountered during random walks within a network.

Ever since its conception, association cortex has been thought to play an important role in integrating information across sensory modalities and in the etiology of neurological syndromes^39^. The localization of gliomas to association cortex provides support for the “tethering hypothesis”, or the notion that the association cortices lack developmental stability compared to the more evolutionarily conserved primary cortices, introducing vulnerability for neuropsychiatric conditions^10,40^. Past work has implicated the tethering hypothesis within the context of psychiatric diseases such as autism and schizophrenia^41,42^. The application of this idea to gliomagenesis suggests that the extensive scaling of association cortex during hominid evolution, an event purportedly responsible for human-unique cognition^40^, may also have introduced risk for brain cancer. This idea is consistent with the observation that genes likely involved in cortical scaling (e.g. neurodevelopmental genes guiding proliferation of neural and glial cells) are affected in glioma^43^.

Early work on gliomagenesis hypothesized that mature glial cells were the cells-of-origin for adult glioma. However, it was soon recognized that the cell-of-origin most likely maintains pluripotency after development, since such cells require fewer mutations to become cancerous^22^. Following recent evidence of NSCs in the subventricular zone and dentate gyrus of the hippocampus of adults, there is an emerging consensus that stem-like cells (including OPCs) could be the cells-of-origin for glioma^21,22^. Recent work has provided strong evidence that some IDH-wildtype glioblastomas originate from stem cells in the subventricular zone^44^. Lee and colleagues demonstrated that for a majority of their glioblastoma patients, the unaffected subventricular zone carried low-level driver mutations, which were present to greater extent in the tumor. Our findings complement this research by establishing that gliomas in general are more highly concentrated in regions enriched with NSCs.

OPCs have also been hypothesized to represent cells-of-origin for glioma. Evidence for this idea comes from studies demonstrating that some gliomas express OPC genetic markers^45,46^, and that OPCs can be experimentally manipulated into becoming cancer stem cells^47,48^. OPCs comprise the majority of dividing cells in the adult brain and are distributed broadly throughout the subventricular zone, white matter, and gray matter^21,26^. We estimated this distribution by quantifying normative expression levels of OPC genetic markers across the human brain, and found that it significantly correlated with glioma frequency. While this result aligns nicely with prior work, estimates of OPC distribution relied on combining data from two independent transcriptomic studies of post-mortem human brains^27,49^. While this approach has been validated for determining the brain-wide distribution of other canonical cell types^28^, our result should be confirmed once reliable estimates of OPC patterning become available.

Normal cells can become malignant through a series of somatic mutations which disable tumor suppressors and activate drivers of cell proliferation^50^. To determine the genetic alterations involved in oncogenesis, much research has focused on identifying molecular genetic differences between tumor cells and matched healthy tissue^51,52^. Here, we took an alternative approach and investigated transcriptomic differences between healthy regions where tumors tend to occur versus healthy regions where tumors are uncommon. As expected, this approach recapitulated prior research into glioma genetics, in that genes which drive gliomagenesis appeared to be upregulated among the healthy transcriptomic correlates of glioma distribution. Gene ontology revealed that the genes driving PLS1 (the component responsible for most of the covariance between transcription and glioma distribution) were most strongly associated with chromatin organization, a process perturbed by IDH mutations and critically involved in the pathogenesis of glioma^31,53^. In addition, our approach also revealed novel findings, such as the carcinogenic vulnerability of healthy brain regions enriched with genes coordinating synaptic signaling and metabolic activity. These findings complement our connectomic results, providing more evidence for the idea that metabolically demanding brain regions crucial for brain-wide communication are susceptible to oncogenesis.

The goal of this study was to examine brain regions generally implicated in adult glioma. However, adult glioma is a heterogeneous phenomenon, comprising tumors of differing genetic etiologies and morphologies. It is known that different types of glioma tend to localize to different brain regions^4,23,24,29^. Therefore, the exact composition of patients (e.g. proportion of high-grade to low-grade glioma patients) within our sample could influence the results. To address this concern, we replicated our results with subgroups of varying proportions of high-grade to low-grade glioma patients and demonstrated that our results are robust to changes to the composition of the sample. However, we did not have access to the molecular genetic characterization of the tumors in our sample, limiting our ability to determine the effect of tumor genotype on the results. Examining glioma subtypes separately could illuminate the network, cellular, and transcriptomic correlates which distinguish localization patterns of different types of glioma. Such work could be useful for developing scientifically informed priors for tumor diagnosis before biopsy, so this question is of both scientific and clinical interest.

## Conclusion

Gaining a better understanding of the mechanisms driving glioma localization patterns could provide a more detailed account of the etiology of the disease and consequently inform treatment targets. We demonstrated that glioma distribution can in part be explained by functional hubness, distribution of stem-like cells, and transcription patterns of genetic determinants of glioma.

These results add to previous literature reporting the vulnerability of hub regions to neurological disease^10,11^, as well as providing support for cancer stem cell theories of glioma^20,22,25^. Our findings highlight the importance of bridging diverse scales of biological organization in the study of oncogenesis.

## Methods

### Tumor Frequency Map

Neuroimaging data of patients with low and high grade gliomas were accessed from the Multimodal Brain Tumor Image Segmentation Challenge 2019 (BraTS: http://braintumorsegmentation.org)^54–56^. T1-weighted contrast-enhanced scans from 259 patients with high-grade glioma and 76 patients with low-grade glioma were segmented by board-certified neuroradiologists, denoting voxels that constituted gadolinium enhancing tumor, non-enhancing core, and peritumoral edema^55^. Segmentation was informed by multimodal imaging, including T1-weighted, post-contrast T1-weighted, T2-weighted, and T2 Fluid Inversion Attenuated Recovery (T2-FLAIR) scans. Scans were acquired at nineteen different institutions (https://www.med.upenn.edu/cbica/brats2019/people.html) with different sequences and protocols. These data were pre-processed through the same pipeline, undergoing linear registration to a common template (SRI24)^57^, resampling at 1mm^3^ isotropic resolution, and removing non-brain tissues from the image^56^.

The minimally pre-processed images were downloaded from the Center for Biomedical Image Computing & Analytics Image Processing Portal (CBICA IPP). Images from each patient were nonlinearly warped to a common template^57^ using Advanced Normalization Tools software (ANTS)^58^, with cost function masking of abnormal brain tissue. The registered masks comprising the gadolinium enhancing tumor and non-enhancing core were taken to represent (and hereafter will be referred to as) the tumor mask.

Tumor masks were concatenated across all 335 patients to create a tumor frequency map, where the value at each voxel denotes the percentage of tumors of the sample that overlapped with that voxel (Figure 1A). Smoothing with a 2 mm full width half maximum (FWHM) Gaussian kernel was applied to the map. An unsmoothed version of this map is shown in Supplementary Information (Supplementary Figure 1). To match genetic data for which most of the samples come from one hemisphere^27^, we mirrored the tumor frequency map to the left hemisphere for the following analyses. Given the large sample size and concordance with other studies^4,59^, this tumor frequency map was interpreted as representing general glioma spatial distribution.

### Tumor Frequency Parcellation

To quantify tumor frequency by common anatomic subdivisions, we applied to the tumor frequency map an in-house 334 region (with 167 left hemisphere regions) parcellation covering 16 subcortical and 318 neocortical areas. This symmetric parcellation was created by applying a back-tracking algorithm that restricts the parcel size to 500 mm^2^ with the Desikan-Killany atlas boundaries as starting points^60^. Although this parcellation was gray matter based, parcels were extended 4 mm into the white matter to capture tumor frequency in adjacent white matter regions. Tumor frequency for a parcel was calculated by averaging the voxel value (representing percent tumor overlap) of the mirrored tumor frequency map within each left hemisphere parcel.

### Internal Replicability

The internal replicability of our tumor frequency map was tested by correlating tumor frequency maps derived from randomly assigned, nonoverlapping cohorts of 168 and 167 patients (Groups 1 and 2).

Simultaneously, we tested the generalizability of our results to groups constituted of differing proportions of low grade versus high grade gliomas. The first group (Group 1) had a 50% higher proportion of low grade gliomas (∼34%) as the full cohort (∼23%), whereas the second group (Group 2) was constituted of a 50% lower proportion of low grade glioma patients (∼11%), These tumor frequency maps were constructed with the same processing as that with the full sample (smoothing, mirroring to the left hemisphere, and parcellation). A 95% confidence interval (CI) for the inter-parcel correlation between Group 1 and Group 2 was determined by constructing a distribution of 100 correlation coefficients where different patients were selected for each group.

### Statistical Inference of Brain Map Correspondence

Several analyses in this study involved investigating the spatial correspondence between different imaging derived measures. In general, this was accomplished by calculating the measures at each parcel in the common parcellation scheme, then correlating these measures across parcels for hypothesis testing. However, since the spatial resolution (and thus the number of parcels) of any parcellation scheme is essentially arbitrary, the actual degrees of freedom cannot be estimated. This is aggravated by the spatial autocorrelation of measures among neighboring parcels that violates the assumption of independent observations. This issue has been addressed in past studies by use of what is termed a “spin test”^61–64^. The spin test procedure is described in more detail in the Supplementary Information. In general, it involves comparing the observed inter-parcel correlation between maps of two measures with a distribution of correlations calculated after one of these maps has been spatially permuted in a way that preserves contiguity among parcels.

### Comparison of Glioma Frequency across Canonical Subnetworks

One question of interest was whether gliomas localized to particular brain subnetworks. Seven canonical subnetworks of the brain (https://surfer.nmr.mgh.harvard.edu/fswiki/CorticalParcellation_Yeo2011)^32^ were mapped onto the tumor frequency map. Violin plots were constructed to compare the distribution of nonzero tumor frequency values between voxels belonging to differing canonical subnetworks.

### Functional Connectome

Glioma frequency was compared to regional connectivity (hubness) as quantified by graph theory metrics applied to the functional connectome derived from resting state fMRI data^65,66^ from over 4000 UK BioBank participants (age range 44-78 years; 53% female). The publicly available “dense voxel-wise connectome” of the first UK BioBank cohort (https://www.fmrib.ox.ac.uk/ukbiobank/) corresponds to a 4-D image, where each voxel consists of 1200 principal components derived from a group-level PCA^67^. A correlation between voxels across components gives a close, memory efficient approximation to the correlation of BOLD signal calculated across concatenated timepoints from all individual participants^67^. The aforementioned 334 region in-house parcellation, covering both subcortical and cortical regions, was applied to the voxel-wise connectome. PCA loadings for voxels within each parcel (i.e. region) were averaged (analogous to a mean timeseries) and correlated between parcels to produce the weights of a graph (Figure 2C). Diagonal elements and negative correlations were set to zero. The same parcellation was also applied to the tumor frequency map to quantify tumor frequency within each parcel. Tumor frequency for a parcel represented the average percentage of lesion overlap of voxels within that parcel. The common parcellation allowed for comparison between measures of tumor frequency and functional hubness.

### Graph Theory Metrics of Hubness

Once the weighted healthy connectome had been constructed, we calculated graph theoretical metrics of hubness using the Brain Connectivity Toolbox^68^. In this graph theoretical approach to neuroimaging data, parcels of the brain are conceived as “nodes”, whereas correlations in functional activity between parcels are conceived as the weights of connections between the nodes. Hub metrics derived included: nodal strength (sum of all weighted connections for a particular node), betweenness centrality (fraction of all shortest paths in a network that pass through a certain node), clustering coefficient (average weighted connections of triangular subgraphs associated with a node), local efficiency (inverse of the average shortest path length between a node and every other node), eigenvector centrality (the extent to which a given brain region connects to other regions with higher centrality), participation coefficient (the strength of connections outside of a node’s given module relative to connections within that node’s module), and within-module degree z-score (nodal strength of a node within its module, compared to within-module nodal strengths of each other node in the module). To reduce the impact of community affiliation on participation coefficient and within-module degree z-score, community affiliations were designated based on the maximum spatial overlap of each node with one of the seven canonical subnetworks^32^.

Hub measures were calculated for each of the 334 nodes of the functional connectome. Measures from homotopic nodes were then averaged together, resulting in 167 observations for each subcortical and cortical parcel per hub metric. Many of the hub measures were observed to have a high correlation with nodal strength. Therefore, we screened out hub measures which had a Spearman’s correlation of *ρ* > 0.95 with nodal strength. This led to the removal of clustering coefficient, local efficiency, and eigenvector centrality. While this threshold is arbitrary, the same result was reached with thresholds ranging from 0.65 to 0.99 (Supplementary Figure 2). Spearman’s correlations were calculated between the remaining hub metrics and tumor frequency and were assessed for significance by comparison to spatially contiguous null models, via the spin test (see Supplementary Information).

### Cellular Correlates of Tumor Frequency

To determine whether tumors were more common in regions enriched for NSCs, we assessed tumor frequency within the two parcels of our 334 region parcellation which most closely aligned with the subventricular zone and the dentate gyrus: the caudate and the hippocampus. The average tumor frequency between these two parcels was compared to average tumor frequency between 10000 random pairs of parcels.

To determine if tumors were more common in regions enriched for OPCs, we compared tumor frequency to an expression map of OPC cell class. This expression map was estimated by assessing transcriptional enrichment of OPC genetic markers using a procedure analogous to that previously described^28^. An OPC gene set was derived from a single cell RNA sequencing study performed on adult postmortem cortical tissue^49^ that determined genes with transcription patterns distinguishing cells by canonical cell types, including excitatory and inhibitory neurons, astrocytes, oligodendrocytes, and OPCs. The set of 132 genes that distinguished OPCs from other canonical cell classes across the cortex was downloaded from previously published material ^49^. Next, we determined parcels where the OPC gene set was upregulated in the adult brain using the publicly available Allen Human Brain Atlas (AHBA)^27^. The Allen Human Brain Atlas catalogues postmortem gene expression from six individuals (ages 24 to 57 years old; five males and one female) at a variety of brain locations. Transcription patterns of 20,647 genes were aligned to the 159 left hemisphere cortical regions in our parcellation, using prior methods^69,70^ with code available for download (https://github.com/RafaelRomeroGarcia/geneExpression_Repository). The resulting 159 × 20,647 regional gene expression matrix was z-scored by parcel. Because the OPC gene set was derived from sequencing performed on cortical brain tissue, we decided to exclude subcortical regions from this part of the analysis.

First, 13 genes in the OPC gene set were not matched to any AHBA probe and were consequently excluded from the analysis. We evaluated the spatial specificity of the remaining 119 OPC genes by comparing their co-expression pattern with 1000 identically-sized sets of randomly chosen genes. OPC genes were filtered out that did not share a positive co-expression pattern with the overall group of genes. Concretely, the 24 genes which had, on average, negative correlations with other genes in the set were removed from the OPC gene set. We estimated OPC distribution by calculating the median regional enrichment of the filtered OPC gene set across cortical parcels. OPC distribution across 159 cortical parcels was then correlated with tumor frequency and tested for significance using the spin test.

### Aligning Tumor Frequency Map with the Allen Human Brain Atlas

Next, we compared tumor frequency with postmortem gene expression from the Allen Human Brain Atlas (http://human.brain-map.org/)^27^. Pre-processing of the AHBA data followed a similar pipeline to previous work from our group^69,70^ and is described in more detail in the Supplementary Information. Transcription levels for 20647 genes across 2748 sample locations were related to tumor frequency at each sample location using partial least squares (PLS) regression. Tumor frequency values were aligned with sample locations by warping the non-smoothed, non-mirrored tumor frequency map into the standard stereotactic space of the Montreal Neurological Institute (MNI), a standard brain template for which the locations of the AHBA microarray samples are known. Once in MNI space, a 2 mm FWHM smoothing kernel was applied to the map and the map was mirrored to the left hemisphere. Sample locations from the AHBA that were located in the right hemisphere were also mirrored to their homotopic voxel in the left hemisphere. This alignment resulted in a 2748 (samples) by 20647 (genes) expression matrix and in a vector of 2748 elements representing tumor frequency values matched to each sample’s MNI coordinates (Figure 4 A,B). Tumor frequency values were square rooted to reduce the skewness of the tumor frequency distribution (Figure 1B). Gene expression values were Z scored for each gene. To test the robustness of the findings, the analyses below were repeated using tumor frequency maps derived from Group 1 and Group 2.

### Transcriptomic Correlates of Tumor Frequency

PLS regression was used to relate spatial transcription patterns of 20647 genes with the spatial distribution of glioma. PLS regression involves projecting a predictor (X) and a response (y) matrix into a space where linear combinations of X explain the maximum amount of variance in y. We chose to focus on the first two components from PLS (PLS 1 and PLS2) as the subsequent components explained a proportion of variance indistinguishable from one another (Supplementary Figure 3A). Statistical significance of the PLS model was tested via permutation testing, by comparing the percent variance explained in the original model to a distribution of 1000 models where the sample labels mapping X to y were randomly shuffled. Significance of each PLS coefficient was tested via bootstrapping with 1000 iterations, resulting in two Z statistics for each gene, one for the first PLS component and another for the second PLS component. Genes were ranked by their Z statistics and entered into gene ontology analyses in GOrilla (http://cbl-gorilla.cs.technion.ac.il/), resulting in a hierarchy of biological terms associated with each PLS component, visualized using Revigo^71^. To ensure a data-driven approach, genes with Z statistic values that did not meet the Bonferroni-corrected significance threshold were not excluded from the gene lists.

### Relating PLS Components to Glioma-related Genes

We sought to determine whether either of our PLS components were enriched for genes that are dysregulated in glioma. We collected a list of 20 genes from a recent review^30^ (listed in the Supplementary Information) that are known to be either mutated, amplified, or lost in specific subtypes of glioma. Four of these genes (*PDGFRA1, RIK, RIS*, and *PI3K*) were not matched to any AHBA probe and were accordingly excluded from the analysis.

Similar to the OPC gene list preprocessing, we first confirmed that these genes co-expressed significantly (compared to 10,000 identically sized sets of genes). Next, we filtered out genes with differing co-expression patterns from the group (denoted by negative correlations, on average, with other genes in the set), leading to the exclusion of three genes (*IDH2, MYCN*, and *CIC*). The median rank of the final list of 13 genes was determined among the first and second PLS components and assessed for significance by comparison to median ranks expected by chance.

### Visualization of PLS Components

We were interested in the locations of the samples which drove each PLS component. First, PLS1 and PLS2 loadings were plotted and colored based of the affiliation of the sample with cortex or subcortex. To determine how PLS1 and PLS2 loadings mapped onto cortex, we assigned samples to parcels via a nearest neighbor mapping. Then, the PLS loading of a parcel was represented as the median PLS loading across samples assigned to that parcel. Two parcels were assigned zero samples from nearest neighbor mapping, and these parcels were assigned the mean loading of the group. More samples were mapped to each subcortical parcel (N=8; mean=78.9; SD=52.1) compared to cortical parcels (N=159; mean=13.3; SD=12.5).

### Multivariate Model Combining Connectomic, Cellular, and Genetic Contributions to Tumor Frequency

To determine how different measures of biological contributions to glioma risk interrelated, we developed a multiple linear regression model combining each of the factors we found to be associated with tumor frequency. The model included nodal strength, OPC distribution, PLS1 loadings, and PLS2 loadings. Each of these measures was represented as a 167-dimensional vector, with a value ascribed to each parcel within our parcellation scheme. The dependent variable for the model was the square root of average tumor frequency within each parcel. The square root of tumor frequency was taken to address the skewness of the original tumor frequency values (Figure 1B). The dependent variable and each of the predictors were Z scored, and zeros were assigned to parcels for which no value could be appropriately calculated (e.g. subcortical parcels for OPC distribution, and parcels mapped to zero samples for PLS1 and PLS2 loadings).

First, we constructed a model to determine whether there were any two-way interaction effects between the different scales of biological factors. Nodal strength represented “connectomic factors”, OPC distribution represented “cellular factors”, and PLS1 and PLS2 loadings represented “genetic factors”. This model had the following form:

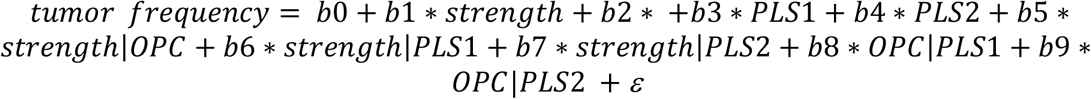

This model revealed no significant interactions effects between different biological factors. Therefore, we constructed a second model with no interaction terms, of the form:

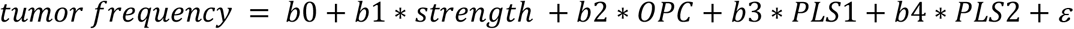

After determining the percentage of explained variance in tumor frequency from these predictors, we explored the individual contribution of each variable by calculating the square of the partial correlation between that variable and tumor frequency. Significance of the explained variance was assessed by comparison to the distribution of explained variances between the variable and 10000 permuted, spatially contiguous, null models of tumor frequency.

## Data Availability

This work made use of publicly-accessible datasets from the Brain Tumor Segmentation Challenge, UK BioBank, and Allen Institute for Brain Science

http://braintumorsegmentation.org/

https://www.fmrib.ox.ac.uk/ukbiobank/

http://human.brain-map.org/

## Acknowledgements

We thank the Brain Tumor Segmentation Challenge (BraTS) for access to the MRI scans and lesion masks used in the study, as well as the patients who participated in that project. This research also relied on imaging data from UK BioBank and genetic data from the Allen Brain Institute. A.S.M. was funded by a Gates Cambridge Scholarship. R.R.G. was funded by a Guarantors of Brain fellowship. Data were stored and processed on the High Performance Hub for Clinical Informatics (HPHI) platform, funded by a Medical Research Council (MRC) infrastructure award (MR/M009041/1).

## Author Contributions

A.S.M., R.R.G., and M.G.H conceived of the research idea. A.S.M. performed the analyses, with R.R.G., M.G.H, and J.S. supervising the work. A.S.M. drafted the article. All authors discussed the results and commented on the manuscript.

## Ethics Statement

The research described in this manuscript used anonymized data made publicly available via the International Brain Tumor Segmentation (BraTS) challenge. All other data considered here had similarly been anonymized before being made publicly accessible.

## Supplemental Information

### Supplementary Methods

#### Spin Test Methodology

The “spin test” involves comparing the observed inter-parcel correlation between maps of two measures with a distribution of the correlations calculated after one of these maps has been spatially permuted in a way that preserves contiguity between brain regions. Spatial permutation was accomplished by projecting the centroid coordinates for each parcel onto an inflation of the pial surface as a sphere^1^, applying a random rotation to that sphere, and then projecting the new coordinates back onto the pial surface and assigning them to the nearest centroid coordinates of the original parcellation. The result is a shuffled parcellation where most parcels remain contiguous.

Past studies using the spin test have focused on comparisons between cortical brain maps. However, subcortical regions were also of interest in this study. Subcortical regions cannot be projected onto the inflated spherical pial surface, so an alternative approach was needed. We incorporated the subcortex into our null models by shuffling the eight subcortical regions with respect to one another, whereas the cortical regions were shuffled using the spin test.

After each spin permutation, two correlations were calculated; one between measures estimated from parcels in their original configuration and the other in its permuted configuration, and vice versa. These two correlations were averaged to form one of the 10000 values forming a null distribution to which the observed correlation was compared to determine statistical significance, as the proportion of null correlations greater than the observed correlation (i.e. *P*_spin_).

#### AHBA Preprocessing

Custom microarrays were used to measure the expression of all genes in the genome in 3702 brain sample locations across cortex, subcortex, and cerebellum ^2^. Pre-processing of these data followed a similar pipeline to previous work from our group ^3,4^. Microarray probes were mapped to genes using the genome assembly hg19 (UCSC GenomeBrowser; http://sourceforge.net/projects/reannotator/) ^5^. In line with criteria from Richiardi and colleagues ^6^, probes were matched to a gene only if there were less than three mismatches between the probe and reference sequence. When a gene matched with multiple probes, the probe with the highest average expression across samples was selected to represent the expression patterns of that gene. A recent study demonstrated the effectiveness of this preprocessing step in increasing the correspondence between microarray and RNA-seq expression ^7^. In total, the expression patterns of 20647 genes across each sample location were evaluated. Samples which were collected from the brain stem and cerebellum were excluded from the analysis, leading to a final number of 2748 samples.

### Supplementary Results

#### Replication of main findings

To determine the robustness of the results, major findings were internally replicated by using sub-cohorts of patients with varying proportions of high- and low-grade glioma (Group 1 and Group 2). Similar to the connectomic results from the full cohort, glioma frequency derived from Group 1 was associated with nodal strength (rho = 0.37; *P*_spin_ = 0.00025), betweenness centrality (rho = 0.48; *P*_spin_ = 0.0002), and participation coefficient (rho = 0.34; *P*_spin_ = 0.0042), but not Z-score modularity (rho = 0.033; *P*_spin_ = 0.33), while glioma frequency derived from Group 2 was associated with nodal strength (rho = 0.29; *P*_spin_ = 0.00033), betweenness centrality (rho = 0.51; *P*_spin_ = 0.0002), and participation coefficient (rho = 0.25; *P*_spin_ = 0.027), but not Z-score modularity (rho = 0.072; *P*_spin_ = 0.19). The association between OPC distribution and glioma frequency was also internally replicated: Group 1: rho = 0.41; *P*_spin_ = 0.0005; Group 2: rho = 0.46; *P*_spin_ = 0.0001. Finally, PLS1 and PLS2 genes lists from the full cohort correlated with gene lists from Group 1 at rho = 0.991 and rho = 0.989 respectively, and with Group 2 at rho = 0.992 and rho = 0.990. PLS1 and PLS2 gene lists from Group 1 and Group 2 correlated with one another at rho = 0.967 and rho = 0.958.

**Supplementary Table 1.** Tumor frequency percentage values at each parcel.

| **Parcel Names** | **% Tumor Frequency** | **Parcel Names (cont.)** | **% Tumor Frequency (cont.)** |
| --- | --- | --- | --- |
| Thalamus-Proper | 3.7 | pericalcarine_part3 | 0.0 |
| Caudate | 8.5 | postcentral_part1 | 0.9 |
| Putamen | 8.8 | postcentral_part2 | 6.9 |
| Pallidum | 6.5 | postcentral_part3 | 1.6 |
| Hippocampus | 9.0 | postcentral_part4 | 2.6 |
| Amygdala | 8.5 | postcentral_part5 | 2.8 |
| Accumbens-area | 4.9 | postcentral_part6 | 1.8 |
| VentralDC | 2.7 | postcentral_part7 | 1.7 |
| bankssts_part1 | 8.0 | postcentral_part8 | 1.9 |
| bankssts_part2 | 12.5 | posteriorcingulate_part1 | 3.5 |
| caudalanteriorcingulate_part1 | 10.8 | posteriorcingulate_part2 | 4.0 |
| caudalmiddlefrontal_part1 | 6.0 | precentral_part1 | 1.3 |
| caudalmiddlefrontal_part2 | 6.1 | precentral_part2 | 6.3 |
| caudalmiddlefrontal_part3 | 3.3 | precentral_part3 | 1.1 |
| caudalmiddlefrontal_part4 | 4.3 | precentral_part4 | 4.3 |
| cuneus_part1 | 0.1 | precentral_part5 | 1.4 |
| cuneus_part2 | 1.5 | precentral_part6 | 3.4 |
| cuneus_part3 | 0.3 | precentral_part7 | 2.4 |
| entorhinal_part1 | 6.7 | precentral_part8 | 2.5 |
| fusiform_part1 | 4.2 | precentral_part9 | 3.5 |
| fusiform_part2 | 6.3 | precuneus_part1 | 2.2 |
| fusiform_part3 | 1.0 | precuneus_part2 | 3.9 |
| fusiform_part4 | 9.4 | precuneus_part3 | 4.4 |
| fusiform_part5 | 3.4 | precuneus_part4 | 2.1 |
| fusiform_part6 | 6.8 | precuneus_part5 | 3.3 |
| inferiorparietal_part1 | 2.6 | precuneus_part6 | 2.3 |
| inferiorparietal_part2 | 8.1 | precuneus_part7 | 3.7 |
| inferiorparietal_part3 | 3.8 | rostralanteriorcingulate_part1 | 7.5 |
| inferiorparietal_part4 | 6.4 | rostralmiddlefrontal_part1 | 5.0 |
| inferiorparietal_part5 | 4.4 | rostralmiddlefrontal_part2 | 5.3 |
| inferiorparietal_part6 | 4.2 | rostralmiddlefrontal_part3 | 2.3 |
| inferiorparietal_part7 | 4.4 | rostralmiddlefrontal_part4 | 5.9 |
| inferiorparietal_part8 | 3.3 | rostralmiddlefrontal_part5 | 3.6 |
| inferiorparietal_part9 | 3.6 | rostralmiddlefrontal_part6 | 6.0 |
| inferiortemporal_part1 | 5.2 | rostralmiddlefrontal_part7 | 4.6 |
| inferiortemporal_part2 | 3.5 | rostralmiddlefrontal_part8 | 7.3 |
| inferiortemporal_part3 | 8.4 | rostralmiddlefrontal_part9 | 5.1 |
| inferiortemporal_part4 | 7.7 | rostralmiddlefrontal_part10 | 5.5 |
| inferiortemporal_part5 | 4.3 | rostralmiddlefrontal_part11 | 5.4 |
| inferiortemporal_part6 | 5.9 | superiorfrontal_part1 | 1.5 |
| isthmuscingulate_part1 | 2.7 | superiorfrontal_part2 | 2.9 |
| isthmuscingulate_part2 | 4.3 | superiorfrontal_part3 | 3.1 |
| lateraloccipital_part1 | 1.4 | superiorfrontal_part4 | 4.0 |
| lateraloccipital_part2 | 4.7 | superiorfrontal_part5 | 2.4 |
| lateraloccipital_part3 | 3.2 | superiorfrontal_part6 | 6.6 |
| lateraloccipital_part4 | 0.3 | superiorfrontal_part7 | 3.0 |
| lateraloccipital_part5 | 0.8 | superiorfrontal_part8 | 7.9 |
| lateraloccipital_part6 | 2.7 | superiorfrontal_part9 | 4.5 |
| lateraloccipital_part7 | 1.2 | superiorfrontal_part10 | 5.2 |
| lateraloccipital_part8 | 0.3 | superiorfrontal_part11 | 4.8 |
| lateraloccipital_part9 | 0.8 | superiorfrontal_part12 | 6.1 |
| lateralorbitofrontal_part1 | 3.3 | superiorfrontal_part13 | 6.1 |
| lateralorbitofrontal_part2 | 3.7 | superiorparietal_part1 | 2.7 |
| lateralorbitofrontal_part3 | 5.0 | superiorparietal_part2 | 3.3 |
| lateralorbitofrontal_part4 | 3.3 | superiorparietal_part3 | 5.7 |
| lateralorbitofrontal_part5 | 6.6 | superiorparietal_part4 | 2.8 |
| lingual_part1 | 2.5 | superiorparietal_part5 | 0.7 |
| lingual_part2 | 0.0 | superiorparietal_part6 | 1.7 |
| lingual_part3 | 2.1 | superiorparietal_part7 | 5.4 |
| lingual_part4 | 0.0 | superiorparietal_part8 | 3.5 |
| lingual_part5 | 1.4 | superiorparietal_part9 | 9.1 |
| lingual_part6 | 0.1 | superiorparietal_part10 | 6.3 |
| medialorbitofrontal_part1 | 2.7 | superiortemporal_part1 | 6.9 |
| medialorbitofrontal_part2 | 3.8 | superiortemporal_part2 | 7.1 |
| medialorbitofrontal_part3 | 2.3 | superiortemporal_part3 | 6.2 |
| medialorbitofrontal_part4 | 4.1 | superiortemporal_part4 | 8.5 |
| middletemporal_part1 | 4.1 | superiortemporal_part5 | 9.4 |
| middletemporal_part2 | 5.1 | superiortemporal_part6 | 12.5 |
| middletemporal_part3 | 6.5 | superiortemporal_part7 | 13.0 |
| middletemporal_part4 | 10.9 | supramarginal_part1 | 9.4 |
| middletemporal_part5 | 8.4 | supramarginal_part2 | 6.0 |
| middletemporal_part6 | 9.0 | supramarginal_part3 | 5.9 |
| parahippocampal_part1 | 7.1 | supramarginal_part4 | 1.8 |
| paracentral_part1 | 0.7 | supramarginal_part5 | 3.2 |
| paracentral_part2 | 3.4 | supramarginal_part6 | 2.4 |
| paracentral_part3 | 0.9 | supramarginal_part7 | 2.9 |
| parsopercularis_part1 | 3.6 | frontalpole_part1 | 1.4 |
| parsopercularis_part2 | 7.1 | temporalpole_part1 | 4.5 |
| parsopercularis_part3 | 7.1 | transversetemporal_part1 | 12.5 |
| parsorbitalis_part1 | 2.9 | insula_part1 | 11.7 |
| parstriangularis_part1 | 3.6 | insula_part2 | 10.0 |
| parstriangularis_part2 | 6.8 | insula_part3 | 11.5 |
| pericalcarine_part1 | 0.2 | insula_part4 | 12.7 |
| pericalcarine_part2 | 1.3 |  |  |

**Supplementary Figure 1.**
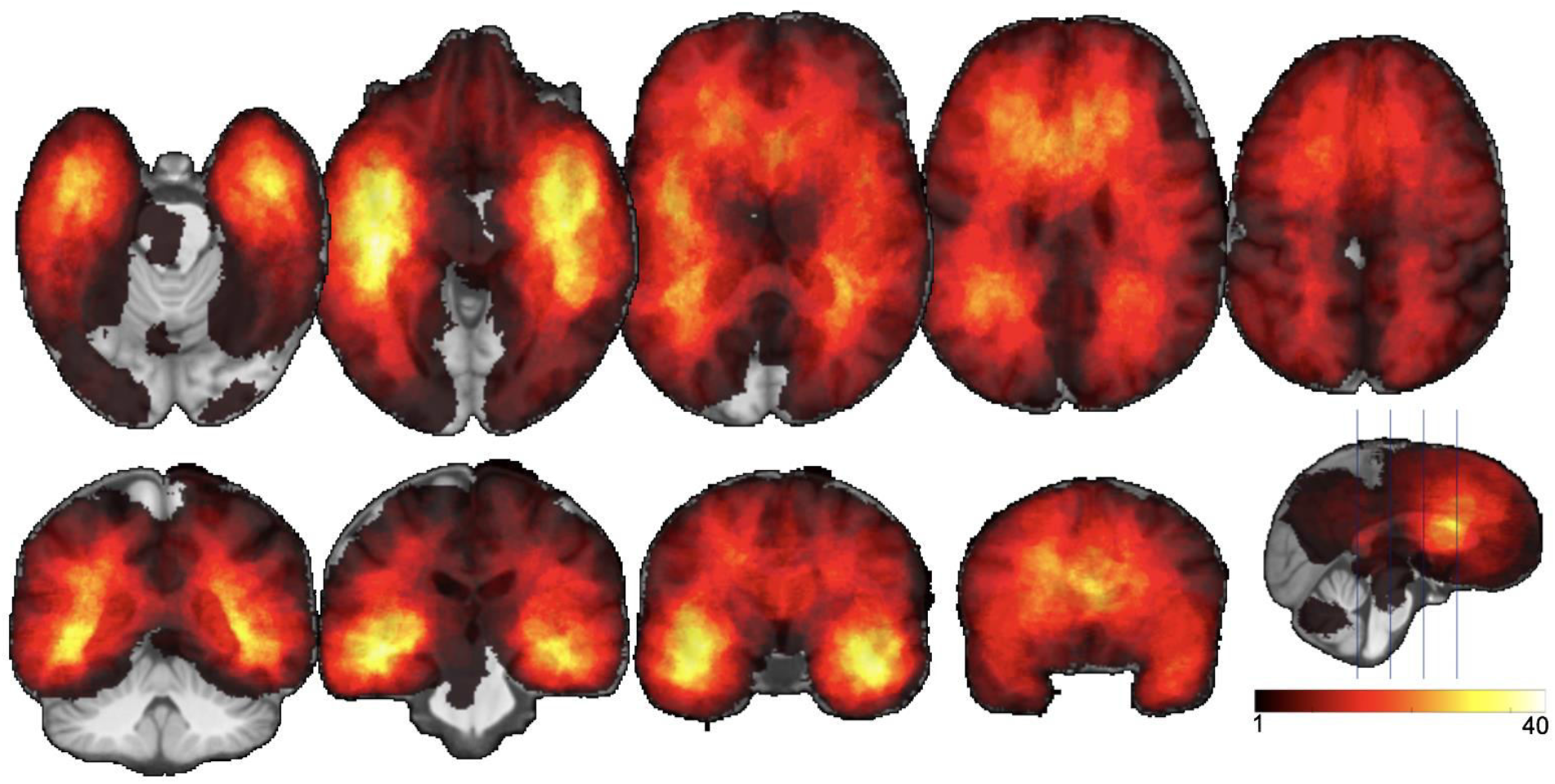
Raw lesion overlap map. Colors indicate the number of lesions (out of a total of 335) overlapping with the associated voxel.

**Supplementary Figure 2.**
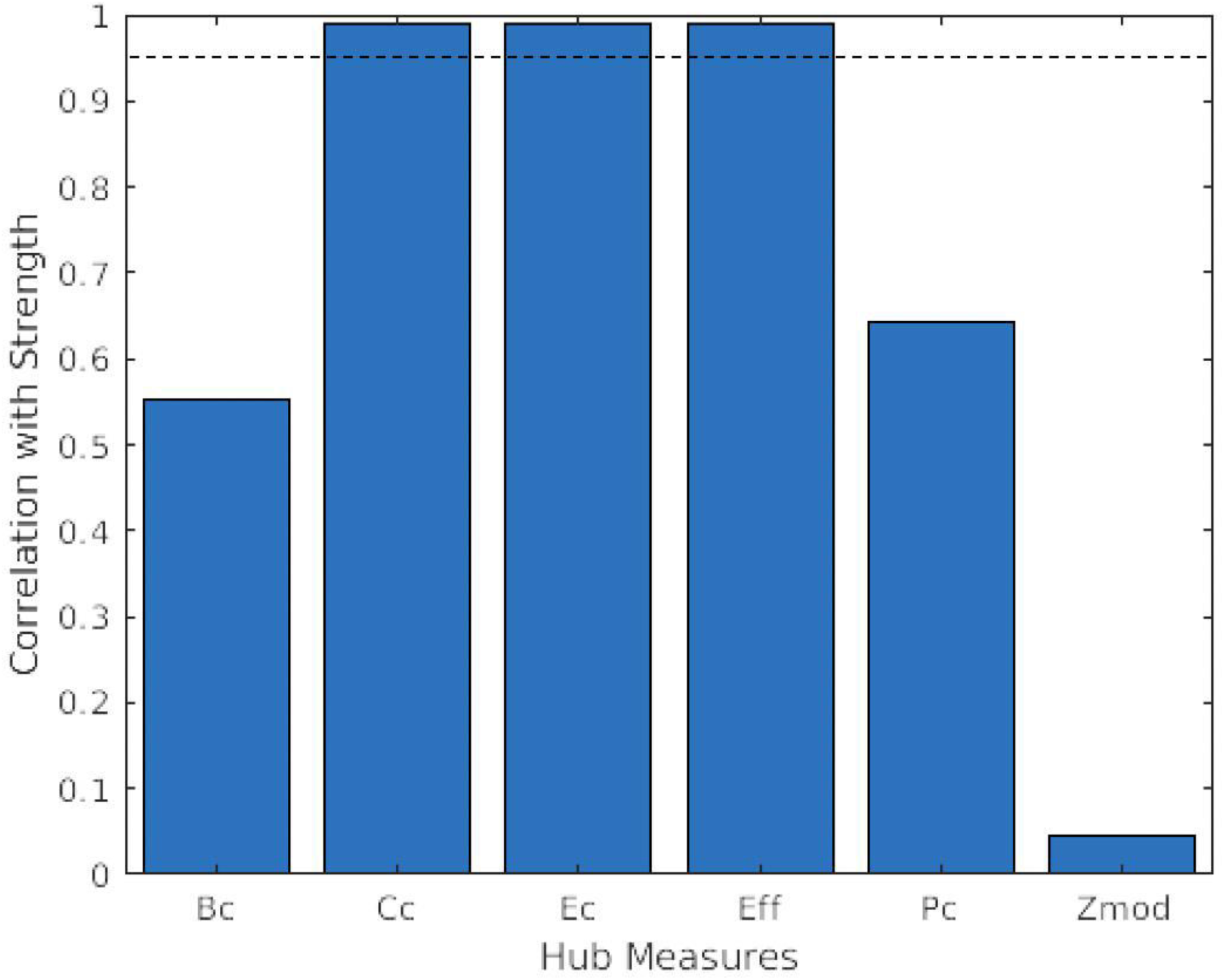
Rank correlations between nodal strength and other graph theoretical metrics of hubness. Graph theoretical metrics with a correlation higher than the dotted line (rho=0.95) were screened from further analyses. While this threshold was arbitrary, the same measures would be screened across a variety of similar thresholds.

**Supplementary Figure 3.**
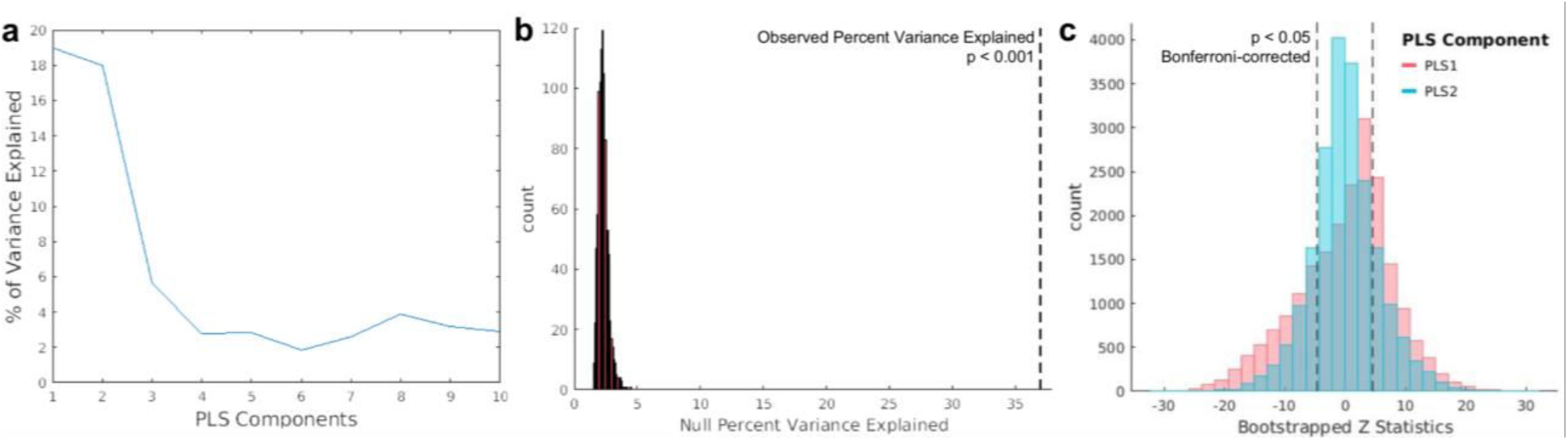
PLS regression analyses relating gene expression with glioma frequency. A. Scree plot demonstrating percentage of variance explained by each subsequent PLS component. B. Percentage of variance explained across 1000 null models where the mapping of glioma frequency to gene expression is randomized, compared to the percent explained variance in the observed model, indicated by the dotted line. C. Distribution of bootstrapped Z statistics for each gene, corresponding to PLS1 and PLS2. Positive and negative Bonferroni-corrected significance thresholds are indicated by the dotted lines. These thresholds were not applied to the PLS gene lists.

## References

1. Jeremic, B. et al. Influence of extent of surgery and tumor location on treatment outcome of patients with glioblastoma multiforme treated with combined modality approach. J. Neurooncol. (1994). doi:10.1007/BF01052902

2. Sagberg, L. M. et al. Brain atlas for assessing the impact of tumor location on perioperative quality of life in patients with high-grade glioma: A prospective population-based cohort study. NeuroImage Clin. (2019). doi:10.1016/j.nicl.2019.101658

3. Bailey, P. & Cushing, H. A Classification of the Tumors of the Glioma Group on a Histogenetic Basis with a Correlated Study of Prognosis. Arch. Neurol. Psychiatry (1926). doi:10.1001/jama.1926.02680040056039

4. Duffau, H. & Capelle, L. Preferential brain locations of low-grade gliomas. Cancer b(2004). doi:10.1002/cncr.20297

5. Duffau, H. Diffuse low-grade gliomas in adults. Diffuse Low-Grade Gliomas in Adults (2017). doi:10.1007/978-3-319-55466-2

6. Seung, S. Connectome: how the brain’s wiring makes us who we are. (Houghton Mifflin Harcourt, 2012).

7. Bullmore, E. & Sporns, O. Complex brain networks: Graph theoretical analysis of structural and functional systems. Nature Reviews Neuroscience (2009). doi:10.1038/nrn2575

8. Bassett, D. S. & Bullmore, E. Small-world brain networks. Neuroscientist (2006). doi:10.1177/1073858406293182

9. van den Heuvel, M. P. & Sporns, O. Network hubs in the human brain. Trends in Cognitive Sciences (2013). doi:10.1016/j.tics.2013.09.012

10. Bullmore, E. & Sporns, O. The economy of brain network organization. Nature Reviews Neuroscience (2012). doi:10.1038/nrn3214

11. Crossley, N. A. et al. The hubs of the human connectome are generally implicated in the anatomy of brain disorders. Brain (2014). doi:10.1093/brain/awu132

12. Warren, D. E. et al. Network measures predict neuropsychological outcome after brain injury. Proc. Natl. Acad. Sci. U. S. A. (2014). doi:10.1073/pnas.1322173111

13. Aerts, H., Fias, W., Caeyenberghs, K. & Marinazzo, D. Brain networks under attack: Robustness properties and the impact of lesions. Brain (2016). doi:10.1093/brain/aww194

14. Seeley, W. W., Crawford, R. K., Zhou, J., Miller, B. L. & Greicius, M. D. Neurodegenerative Diseases Target Large-Scale Human Brain Networks. Neuron (2009). doi:10.1016/j.neuron.2009.03.024

15. Nijssen, J., Comley, L. H. & Hedlund, E. Motor neuron vulnerability and resistance in amyotrophic lateral sclerosis. Acta Neuropathologica (2017). doi:10.1007/s00401-017-1708-8

16. Buzsáki, G., Geisler, C., Henze, D. A. & Wang, X. J. Interneuron Diversity series: Circuit complexity and axon wiring economy of cortical interneurons. Trends in Neurosciences (2004). doi:10.1016/j.tins.2004.02.007

17. Magistretti, P. J. & Allaman, I. A Cellular Perspective on Brain Energy Metabolism and Functional Imaging. Neuron (2015). doi:10.1016/j.neuron.2015.03.035

18. Wodarz, D. Effect of stem cell turnover rates on protection against cancer and aging. J. Theor. Biol. (2007). doi:10.1016/j.jtbi.2006.10.013

19. Rinaldi, M. et al. ROS and brain gliomas: An overview of potential and innovative therapeutic strategies. International Journal of Molecular Sciences (2016). doi:10.3390/ijms17060984

20. Visvader, J. E. Cells of origin in cancer. Nature (2011). doi:10.1038/nature09781

21. Jiang, Y. & Uhrbom, L. On the origin of glioma. Ups. J. Med. Sci. (2012). doi:10.3109/03009734.2012.658976

22. Sanai, N., Alvarez-Buylla, A. & Berger, M. S. Neural Stem Cells and the Origin of Gliomas. N. Engl. J. Med. (2005). doi:10.1056/nejmra043666

23. Zlatescu, M. C. et al. Tumor location and growth pattern correlate with genetic signature in oligodendroglial neoplasms. Cancer Res. (2001).

24. Mueller, W. et al. Genetic signature of oligoastrocytomas correlates with tumor location and denotes distinct molecular subsets. Am. J. Pathol. (2002). doi:10.1016/S0002-9440(10)64183-1

25. Ma, D. K., Bonaguidi, M. A., Ming, G. L. & Song, H. Adult neural stem cells in the mammalian central nervous system. Cell Research (2009). doi:10.1038/cr.2009.56

26. Hughes, E. G., Kang, S. H., Fukaya, M. & Bergles, D. E. Oligodendrocyte progenitors balance growth with self-repulsion to achieve homeostasis in the adult brain. Nat. Neurosci. (2013). doi:10.1038/nn.3390

27. Hawrylycz, M. J. et al. An anatomically comprehensive atlas of the adult human brain transcriptome. Nature (2012). doi:10.1038/nature11405

28. Seidlitz, J. et al. Transcriptomic and Cellular Decoding of Regional Brain Vulnerability to Neurodevelopmental Disorders. bioRxiv (2019). doi:10.1101/573279

29. Tejada Neyra, M. A. et al. Voxel-wise radiogenomic mapping of tumor location with key molecular alterations in patients with glioma. Neuro. Oncol. (2018). doi:10.1093/neuonc/noy134

30. Molinaro, A. M., Taylor, J. W., Wiencke, J. K. & Wrensch, M. R. Genetic and molecular epidemiology of adult diffuse glioma. Nat. Rev. Neurol. 15, (2019).

31. Reifenberger, G., Wirsching, H. G., Knobbe-Thomsen, C. B. & Weller, M. Advances in the molecular genetics of gliomas-implications for classification and therapy. Nature Reviews Clinical Oncology (2017). doi:10.1038/nrclinonc.2016.204

32. Thomas Yeo, B. T. et al. The organization of the human cerebral cortex estimated by intrinsic functional connectivity. J. Neurophysiol. (2011). doi:10.1152/jn.00338.2011

33. Mufford, M. S. et al. Neuroimaging genomics in psychiatry-a translational approach. Genome Medicine (2017). doi:10.1186/s13073-017-0496-z

34. Henderson, M. X. et al. Spread of α-synuclein pathology through the brain connectome is modulated by selective vulnerability and predicted by network analysis. Nat. Neurosci. (2019). doi:10.1038/s41593-019-0457-5

35. Zhou, J., Gennatas, E. D., Kramer, J. H., Miller, B. L. & Seeley, W. W. Predicting Regional Neurodegeneration from the Healthy Brain Functional Connectome. Neuron (2012). doi:10.1016/j.neuron.2012.03.004

36. Brown, J. A. et al. Patient-Tailored, Connectivity-Based Forecasts of Spreading Brain Atrophy. Neuron (2019). doi:10.1016/j.neuron.2019.08.037

37. Pedersen, P.-H et al. Migratory patterns of lac-z transfected human glioma cells in the rat brain. Int. J. Cancer (1995). doi:10.1002/ijc.2910620620

38. Venkatesh, H. S. et al. Electrical and synaptic integration of glioma into neural circuits. Nature (2019). doi:10.1038/s41586-019-1563-y

39. Mesulam, M. M. Fifty years of disconnexion syndromes and the Geschwind legacy. Brain (2015). doi:10.1093/brain/awv198

40. Buckner, R. L. & Krienen, F. M. The evolution of distributed association networks in the human brain. Trends in Cognitive Sciences (2013). doi:10.1016/j.tics.2013.09.017

41. van den Heuvel, M. P. et al. Evolutionary modifications in human brain connectivity associated with schizophrenia. Brain (2019). doi:10.1093/brain/awz330

42. Wei, Y. et al. Genetic mapping and evolutionary analysis of human-expanded cognitive networks. Nat. Commun. (2019). doi:10.1038/s41467-019-12764-8

43. Puchalski, R. B. et al. An anatomic transcriptional atlas of human glioblastoma. Science (80-.). (2018). doi:10.1126/science.aaf2666

44. Lee, J. H. et al. Human glioblastoma arises from subventricular zone cells with low-level driver mutations. Nature (2018). doi:10.1038/s41586-018-0389-3

45. Shoshan, Y. et al. Expression of oligodendrocyte progenitor cell antigens by gliomas: Implications for the histogenesis of brain tumors. Proc. Natl. Acad. Sci. U. S. A. (1999). doi:10.1073/pnas.96.18.10361

46. Verhaak, R. G. W. et al. Integrated Genomic Analysis Identifies Clinically Relevant Subtypes of Glioblastoma Characterized by Abnormalities in PDGFRA, IDH1, EGFR, and NF1. Cancer Cell (2010). doi:10.1016/j.ccr.2009.12.020

47. Kondo, T. & Raff, M. Oligodendrocyte precursor cells reprogrammed to become multipotential CNS stem cells. Science (80-.). (2000). doi:10.1126/science.289.5485.1754

48. Liu, C. et al. Mosaic analysis with double markers reveals tumor cell of origin in glioma. Cell (2011). doi:10.1016/j.cell.2011.06.014

49. Lake, B. B. et al. Integrative single-cell analysis of transcriptional and epigenetic states in the human adult brain. Nat. Biotechnol. (2018). doi:10.1038/nbt.4038

50. Mukherjee, S. The Emperor of All Maladies: A Biography of Cancer. J. Postgrad. Med. Educ. Res. (2012). doi:10.5005/jp-journals-10028-1025

51. Tang, J., He, D., Yang, P., He, J. & Zhang, Y. Genome-wide expression profiling of glioblastoma using a large combined cohort. Sci. Rep. (2018). doi:10.1038/s41598-018-33323-z

52. McLendon, R. et al. Comprehensive genomic characterization defines human glioblastoma genes and core pathways. Nature (2008). doi:10.1038/nature07385

53. Suzuki, H. et al. Mutational landscape and clonal architecture in grade II and III gliomas. Nat. Genet. (2015). doi:10.1038/ng.3273

54. Menze, B. H. et al. The Multimodal Brain Tumor Image Segmentation Benchmark (BRATS). IEEE Trans. Med. Imaging (2015). doi:10.1109/TMI.2014.2377694

55. Bakas, S. et al. Advancing The Cancer Genome Atlas glioma MRI collections with expert segmentation labels and radiomic features. Sci. Data (2017). doi:10.1038/sdata.2017.117

56. Bakas, S. et al. Identifying the Best Machine Learning Algorithms for Brain Tumor Segmentation, Progression Assessment, and Overall Survival Prediction in the BRATS Challenge. arXiv (2018).

57. Rohlfing, T., Zahr, N. M., Sullivan, E. V. & Pfefferbaum, A. The SRI24 multichannel atlas of normal adult human brain structure. Hum. Brain Mapp. (2010). doi:10.1002/hbm.20906

58. Avants, B. B., Tustison, N. & Song, G. Advanced Normalization Tools (ANTS). Insight J. 1–35 (2009). doi:http://hdl.handle.net/10380/3113

59. Larjavaara, S. et al. Incidence of gliomas by anatomic location. Neuro. Oncol. (2007). doi:10.1215/15228517-2007-016

60. Romero-Garcia, R., Atienza, M., Clemmensen, L. H. & Cantero, J. L. Effects of network resolution on topological properties of human neocortex. Neuroimage (2012). doi:10.1016/j.neuroimage.2011.10.086

61. Alexander-Bloch, A., Raznahan, A., Bullmore, E. & Giedd, J. The convergence of maturational change and structural covariance in human cortical networks. J. Neurosci. (2013). doi:10.1523/JNEUROSCI.3554-12.2013

62. Vandekar, S. N. et al. Topologically dissociable patterns of development of the human cerebral cortex. J. Neurosci. (2015). doi:10.1523/JNEUROSCI.3628-14.2015

63. Alexander-Bloch, A. F. et al. On testing for spatial correspondence between maps of human brain structure and function. Neuroimage (2018). doi:10.1016/j.neuroimage.2018.05.070

64. Váša, F. et al. Adolescent tuning of association cortex in human structural brain networks. Cereb. Cortex (2018). doi:10.1093/cercor/bhx249

65. Miller, K. L. et al. Multimodal population brain imaging in the UK Biobank prospective epidemiological study. Nat. Neurosci. (2016). doi:10.1038/nn.4393

66. Alfaro-Almagro, F. et al. Image processing and Quality Control for the first 10,000 brain imaging datasets from UK Biobank. Neuroimage (2018). doi:10.1016/j.neuroimage.2017.10.034

67. Smith, S. M., Hyvärinen, A., Varoquaux, G., Miller, K. L. & Beckmann, C. F. Group-PCA for very large fMRI datasets. Neuroimage (2014). doi:10.1016/j.neuroimage.2014.07.051

68. Rubinov, M. & Sporns, O. Complex network measures of brain connectivity: Uses and interpretations. Neuroimage 52, 1059–1069 (2010).

69. Romero-Garcia, R. et al. Structural covariance networks are coupled to expression of genes enriched in supragranular layers of the human cortex. Neuroimage (2018). doi:10.1016/j.neuroimage.2017.12.060

70. Romero-Garcia, R. et al. Schizotypy-related magnetization of cortex in healthy adolescence is co-located with expression of schizophrenia-related genes. Biol. Psychiatry (2019). doi:10.1016/j.biopsych.2019.12.005

71. Supek, F., Bošnjak, M., škunca, N. & šmuc, T. Revigo summarizes and visualizes long lists of gene ontology terms. PLoS One (2011). doi:10.1371/journal.pone.0021800

## References

1. Fischl, B. FreeSurfer. NeuroImage (2012). doi:10.1016/j.neuroimage.2012.01.021

2. Hawrylycz, M. J. et al. An anatomically comprehensive atlas of the adult human brain transcriptome. Nature (2012). doi:10.1038/nature11405

3. Romero-Garcia, R. et al. Structural covariance networks are coupled to expression of genes enriched in supragranular layers of the human cortex. Neuroimage (2018). doi:10.1016/j.neuroimage.2017.12.060

4. Romero-Garcia, R. et al. Schizotypy-related magnetization of cortex in healthy adolescence is co-located with expression of schizophrenia-related genes. Biol. Psychiatry (2019). doi:10.1016/j.biopsych.2019.12.005

5. Arloth, J., Bader, D. M., Röh, S. & Altmann, A. Re-Annotator: Annotation pipeline for microarray probe sequences. PLoS One (2015). doi:10.1371/journal.pone.0139516

6. Richiardi, J. et al. Correlated gene expression supports synchronous activity in brain networks. Science (80-.). (2015). doi:10.1126/science.1255905

7. Arnatkevičiūtė, A., Fulcher, B. D. & Fornito, A. A practical guide to linking brain-wide gene expression and neuroimaging data. Neuroimage (2019). doi:10.1016/j.neuroimage.2019.01.011

8. Molinaro, A. M., Taylor, J. W., Wiencke, J. K. & Wrensch, M. R. Genetic and molecular epidemiology of adult diffuse glioma. Nat. Rev. Neurol. 15, (2019).

